# Influences on Preventive Health Behaviors After Stroke with Telemonitoring: Patient Perceptions and Practices in an Urban Underserved Population

**DOI:** 10.64898/2026.04.28.26352004

**Authors:** Imama A. Naqvi, Keri Fisher, Kevin Strobino, Adriana Arcia, Clare Bassile, Sapana R. Patel, Ying Kuen Cheung, Joel Stein, Olajide A. Williams, Mitchell S.V. Elkind, Ian M. Kronish, Lori Quinn

**Affiliations:** Department of Neurology, Vagelos College of Physicians and Surgeons, Columbia University, New York, NY; Department of Biobehavioral Sciences, Teachers College, Columbia University, New York, NY; Hahn School of Nursing and Health Science, University of San Diego, San Diego, CA; Department of Rehabilitation and Regenerative Medicine, Vagelos College of Physicians and Surgeons, Columbia University, New York, NY; Department of Psychiatry, Vagelos College of Physicians and Surgeons, Columbia University, New York, NY and the New York State Psychiatric Institute; Department of Biostatistics, Mailman School of Public Health, Columbia University, New York, NY; Department of Rehabilitation Medicine, Weill Cornell Medical College, New York, NY, USA; Department of Epidemiology, Mailman School of Public Health, Columbia University, New York, NY; Department of Medicine, Center for Behavioral Cardiovascular Health, Columbia University, New York, NY

**Author notes:** **Corresponding Author** Imama A. Naqvi, MD, MS, The Neurological Institute of New York, New York City, NY, USA.

**Keywords:** Stroke, Hypertension, Health Behaviors, Disparities, Self-efficacy, Agency

## Abstract

**Background:** Although Blood Pressure (BP) self-management and physical activity (PA) are secondary stroke preventive behaviors, adherence gaps exist. This study explored factors influencing these behaviors after telemonitoring experience among patients in an underserved urban community.

**Methods:** We conducted semi-structured interviews with purposive sampling of patients discharged home after mild stroke who had hypertension and participated in the Telehealth After Stroke Care (TASC) trial. Self-reported short form (SF) surveys included Patient Reported Outcomes Measurement Information System-Physical Function-SF, International Physical Activity Questionnaire-SF, and University of Rhode Island Change Assessment. The first interview assessed knowledge of BP and PA guidelines with perceived barriers, facilitators and BP telemonitoring experience, while the second was after PA monitoring for one month. We performed open (inductive) and social cognitive theory-based (deductive) coding.

**Results:** We included 14 participants: mean age 59 ± 9.6 years; 7 women (50%); 57% Black, 29% Hispanic; 29% ≤ high school education, 43% Medicaid or no insurance. Mean daily step count was 5147 ± 2534. Three themes interpreted included: 1) positive outcome expectations; 2) self-efficacy; and 3) agency. Participants associated BP control with reduced recurrence risk and PA to functional recovery (1) but lacked knowledge of specific targets (2). High self-efficacy individuals (2) used action planning to navigate environmental constraints. Both BP and PA monitoring with feedback facilitated self-regulation, goal setting and problem solving (3).

**Conclusion:** Gaps between knowledge of and participation in health behaviors after stroke persist. Targeting outcome expectations, self-efficacy, and agency through educated training, tailored support, and telemonitored feedback may promote sustained positive health behaviors.

## INTRODUCTION

Recurrent stroke constitutes almost 25% of the nearly 800,000 strokes that occur annually in the US.^1^ While there has been a decrease in recurrent stroke rates, disparities persist.^2^ Black adults are 60% more likely to experience a recurrent stroke within two years than White individuals.^3^ On average, secondary stroke results in poorer outcomes than an incident stroke.^4^

Preventive behavioral strategies such as blood pressure (BP) medication adherence and increased physical activity (PA) can reduce recurrent stroke events by up to 80%.^5^ However, there is a gap between recommendations and guideline uptake by patients and providers. Hypertension is the most important modifiable risk factor for stroke,^6^ yet BP remains poorly controlled in >55% stroke patients..^7–11^ BP control is poorer among Hispanic and Black patients compared to White patients with up to a third reporting low medication adherence.^12^ In addition, higher habitual PA is linked to lower BP.^13^ In fact, even low to moderate intensity PA reduces risk of recurrent events.^14^ Yet, regardless of time since stroke, many survivors spend the majority of their day in sedentary behaviors^15^ and have lower step counts than people of similar ages.^16^

Self-management programs have been created to shift the paradigm from passive education and information sharing to one where individuals play an active role in their disease management.^17^ Among stroke survivors, self-management programs alone have not been found to significantly affect medical risk factors, but can improve lifestyle behavior such as medication adherence.^18^ However, behavioral change strategies such as lifestyle motivational interviewing alone fails to improve the uptake of health-related behaviors.^19,20^ Further, passive use of digital technology through devices to improve health-related behaviors may not be enough to induce lasting change for self-management among stroke survivors.^21,22^ Therein lies a requisite for intentional behavioral change theory-backed application and feasibility testing of active ingredients driving the underlying health behavior change for secondary stroke prevention.^23–25^

Immediately post-stroke, patients face challenges in their transition from the hospital directly back to home.^26,27^ Individual patient-level social drivers such as low health literacy, job and housing insecurity,^28^ together with systemic fragmented transitions of care^29^ leave these patients “floundering”^30^ to self-manage the mounting burden with limited attentional capacity to engage in preventative behaviors. Further, evidence shows that belonging to an urban underserved community with neighborhood crime, unreliable public transportation and limited local resources hampers recovery.^31^ Ultimately, these barriers may take their attention further away from health preventive behaviors for optimal self-management.^27^ Our previously published Telehealth After Stroke Care (TASC) pilot, examined home BP telemonitoring-enhanced versus usual post-acute stroke care in an underserved setting.^32^ It included multidisciplinary team-based care with wireless BP devices and infographics delivered via telehealth after discharge home. We reported significantly improved home systolic BP control (<130/80 mmHg) in the intervention group compared to usual care (76% vs. 25%, p<0.01), with a total difference of 18.4 ± 22 mm Hg between the two groups, suggesting that TASC influenced preventive behaviors including medication adherence and home BP monitoring. The intervention group had better adherence to video visits as compared with usual care,^28^ with high adherence to BP telemonitoring in the intervention group at 91% at one month to 86% at 3 months. (Suppl.) Patient Reported Outcomes (PROs) suggested better adherence to medications with multidisciplinary support.^33^ However, patients’ experiences with TASC have not been evaluated systematically using theory-informed methods.

A theoretical basis for characterizing and designing behavior change interventions is fundamental for successful implementation.^34,35^ Social Cognitive Theory (SCT) could serve as the theoretical framework to define the active ingredients involved in development and maintenance of these secondary stroke prevention behaviors.^36^ SCT was founded on the idea that individuals are agents of their own change; it emphasizes the interaction between cognitive, behavioral, and environmental patterns (influences).^37^ The comprehensive scope of SCT inherently addresses the influence of intrinsic and extrinsic burden of disease and social drivers of health (SDOH). By analyzing the specific barriers and facilitators to health behaviors within this framework we can better understand the cumulative complexity^30^ that arises during stroke transitions of care.

Our primary aim was to understand factors that influence secondary stroke preventive health behaviors from stroke survivors who reside in an underserved urban community. Our secondary aim was to understand the behavioral underpinnings of improved BP outcomes noted from TASC trial components of home BP-telemonitoring with multidisciplinary team-based care in a conceptual model (Interview 1). As it is common for risk factors to co-occur,^38,39^ we also explored PA behaviors (Interview 1) and the experiences of these individuals after PA monitoring (Interview 2). Here we describe factors influencing participation in multiple health-related activities with telemonitoring, including BP (medication adherence and monitoring) and PA (behaviors and monitoring) among these stroke survivors.

## METHODS

### Study Design

Purposive sampling^40^ with critical case criteria^41^ was used for an anticipated adequate but small sample size to capture patient experiences in this qualitative study.^42^ Participants were recruited from the previously published TASC trial.^43^ Eligible study participants included those with a mildly disabling acute ischemic or hemorrhagic stroke discharged home after hospitalization with a diagnosis of hypertension and ability to provide consent. All participants enrolled had presented to the Columbia University Irving Medical Center (CUIMC) Comprehensive Stroke Center in Northern Manhattan that serves a predominantly Hispanic low-income community. Participants were contacted through a phone call to request participation, and interviews were scheduled after remote informed consent. Figure 1 outlines the timeline from patient discharge, enrollment in the TASC trial and study procedures.

**Figure 1.**
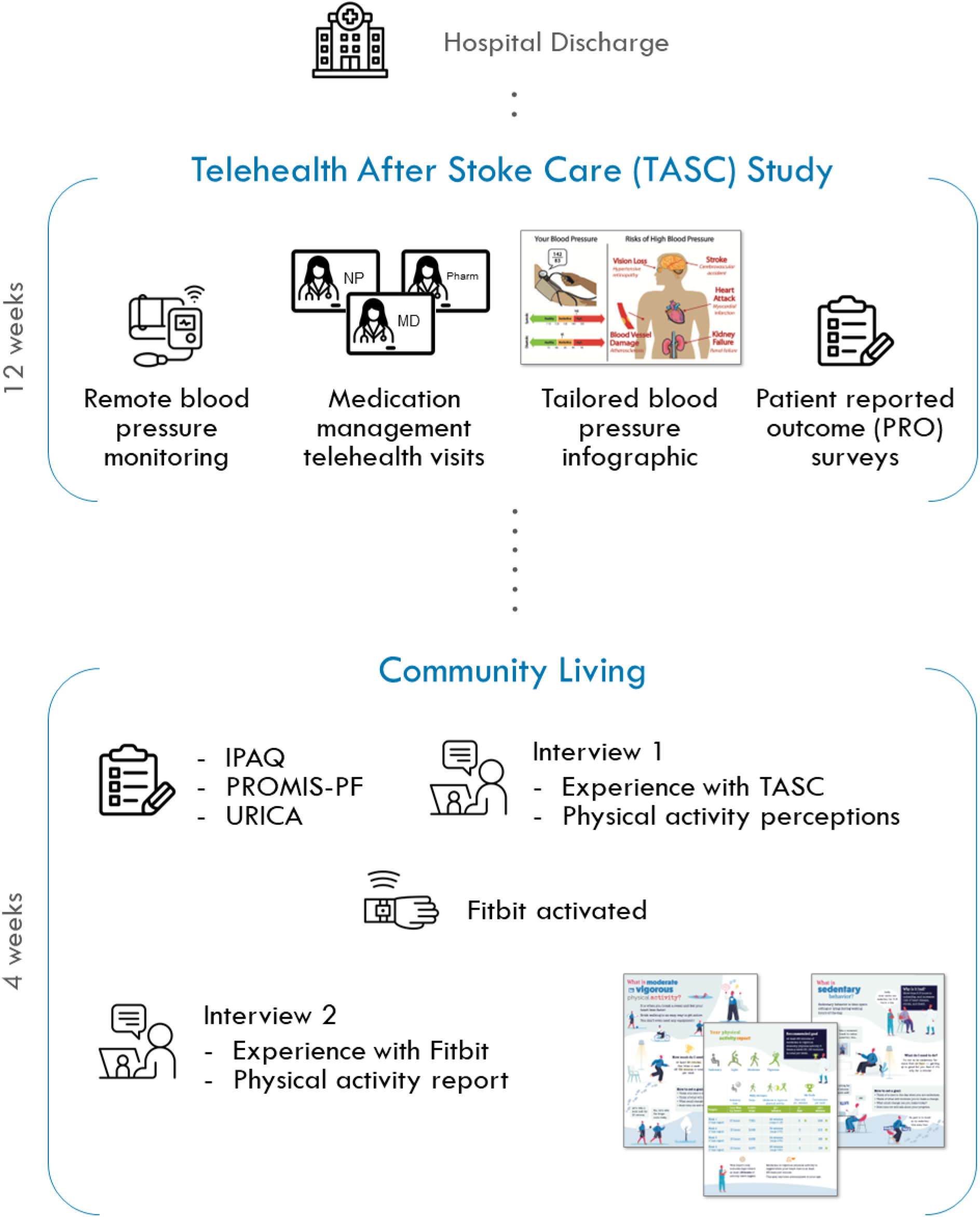
Timeline from hospital discharge to community living and study workflow with mixed methods.

### Patient Interviews

Fourteen of 21 retained TASC intervention participants contacted agreed to enroll. Two semi-structured interviews using a structured interview guide were conducted for each participant. Interviews were conducted at two time-points (Figure 1). Interview 1 occurred one year after TASC completion and was focused on participant experiences in TASC including BP medication use and BP monitoring, and current PA behavior. Interview 2 took place 4 weeks later after wearing a PA monitor (Fitbit Inspire 2, San Francisco, CA). This second interview focused on both the experience of wearing a PA monitor and receiving feedback on activity through a tailored PA infographic using their own data which was shown to them at this interview. It included sedentary time, daily steps, moderate-vigorous physical activity (MVPA) minutes and recommended goals that was shown at the time of this second interview.

The study principal investigator (IAN), a female stroke neurologist with more than 10 years of experience caring for stroke patients conducted interviews with English-speaking participants. The participants were all known to the interviewer from previously participating in TASC. For the one participant that spoke Spanish only, a native Spanish speaker (CC) conducted the interview. All interviews were conducted via HIPAA-compliant Zoom^44^ using a semi-structured interview guide and lasted 30-60 minutes. The audio of the call was recorded using Zoom on an encrypted Columbia University IT-secured device and transcribed using Microsoft Stream.^45^ Spanish transcriptions were translated through Columbia University approved services.

### PA Monitoring

PA monitoring was conducted using Fitbits that have been shown to be an acceptable, feasible, and low-cost way to monitor PA and measure step count after stroke.^46–48^ After completion of the first interview, the study coordinator (CC) sent the programmed Fitbit to the participant. Participants were contacted via phone and/or email by a researcher (KF) once notified by the study coordinator that the activity monitor had been shipped. Participants were instructed to charge it upon receipt. During a follow-up call (within 1 week), the researcher provided instructions on downloading the Fitbit application, signing in with the participant’s assigned username/password, and pairing it. Participants were encouraged to wear the activity monitor for awake and asleep hours. They were contacted if a 3-day gap in Fitbit data was noted during regular Fitabase compliance monitoring. If a participant or proxy was less proficient with technology or the use of smartphone applications, the call was completed over video. (Suppl.).

### Patient Reported Outcomes

Socio-demographics were derived from interviews and/or medical records. Additional study measures were obtained via surveys after enrollment. Surveys were completed online through REDcap or verbally with the study coordinator based on participant comfort with computerized assessments. The PROMIS Physical Function-Short Form (PROMIS-PF) is a patient reported measure of perceived physical function. It is highly correlated with the Stroke Impact Scale-16 (SIS-16;and has excellent internal consistency and a low ceiling effect.^49^ It was scored using a scoring algorithm for REDCap provided by the HealthMeasures service to produce standardized t-score estimates with a mean of 50 and a standard deviation of 10, where higher scores indicate higher levels of the target trait (e.g., better physical function).^50^

The International Physical Activity Questionnaire Short Form (IPAQ-SF) yields valid and reliable measure used to assess self-reported PA among stroke survivors.^51^ Participants were asked to recall their activity over the last seven days and results were used to categorize PA into low (3.3 METs), moderate (4.0 METs), or high (8.0 METs) levels. Scores were calculated by multiplying the average number of hours per day for each type of PA by its corresponding Metabolic Equivalent Task (MET) value.^52^

University of Rhode Island Change Assessment - Exercise (URICA-E2) is a continuous measure of behavioral readiness for exercise.^53^ Participants were asked to report on the stage of readiness to change (precontemplation non-believer, precontemplation believer, contemplation, preparation, action, or maintenance) through which they were passing when changing behavior for exercise.

### Data Analysis Methods

#### PA Data Analysis

Objective data from the activity monitors was aggregated for each participant and analyzed using Fitabase (San Diego, CA).^54^ We used the Fitbit minute-by-minute heart rate output to define a valid wear day as any day (starting and ending at midnight) with 10 hours of continuous heart rate recordings, allowing for an interruption of no more than 90 continuous minutes. If there was a gap of greater than 90 minutes, the 10 hours of continuous data needed to be prior to or after the time gap. Any non-zero value for beats per minute contributed to meeting the heart rate criterion.^55^

A week was considered valid if it included a period of seven consecutive days containing at least 4 valid days.^55^ The percentage of valid days (adherence) was calculated by dividing the number of valid days over “wear time”, i.e. the total time participant was using the Fitbit.^56^ Moderate-vigorous physical activity (MVPA) was based on Fitbit “fairly active + very active” minutes, which has been reported as over 3 METS.^57^

#### Qualitative Data Analysis

Deidentified transcripts were cleaned of extraneous utterances by a study researcher (KF). Cleaned versions were uploaded into Dedoose Version 9.0 Software program^58^ for qualitative analysis. The material was in English and did not need to be translated other than from one participant where it was translated from Spanish before analysis. Separate data sets were initially made for interviews 1 (post-TASC BP management and PA behaviors) and 2 (post-PA monitoring feedback).

A hybrid inductive-deductive thematic analysis was conducted. Initial codes were generated inductively from participants narratives, followed by organization into patterns related to constructs from the SCT framework. Codes from interviews 1 and 2 were combined when similar themes were identified under SCT for both BP and PA behaviors and monitoring. Codes then collapsed into categories when representing similar concepts and expanded into subcodes when necessary. Transcripts were reviewed multiple times and memos were used to document emerging patterns and guide interpretative decisions. Seven preliminary themes were identified and refined into three overarching themes that reflect patterns across participants. A data display of select quotes within each theme was created to examine variations in influences of preventive health behaviors (Figure 2). The data display was aligned with SCT constructs to reflect variations across cognitive, behavioral and environmental, and domains.

**Figure 2.**
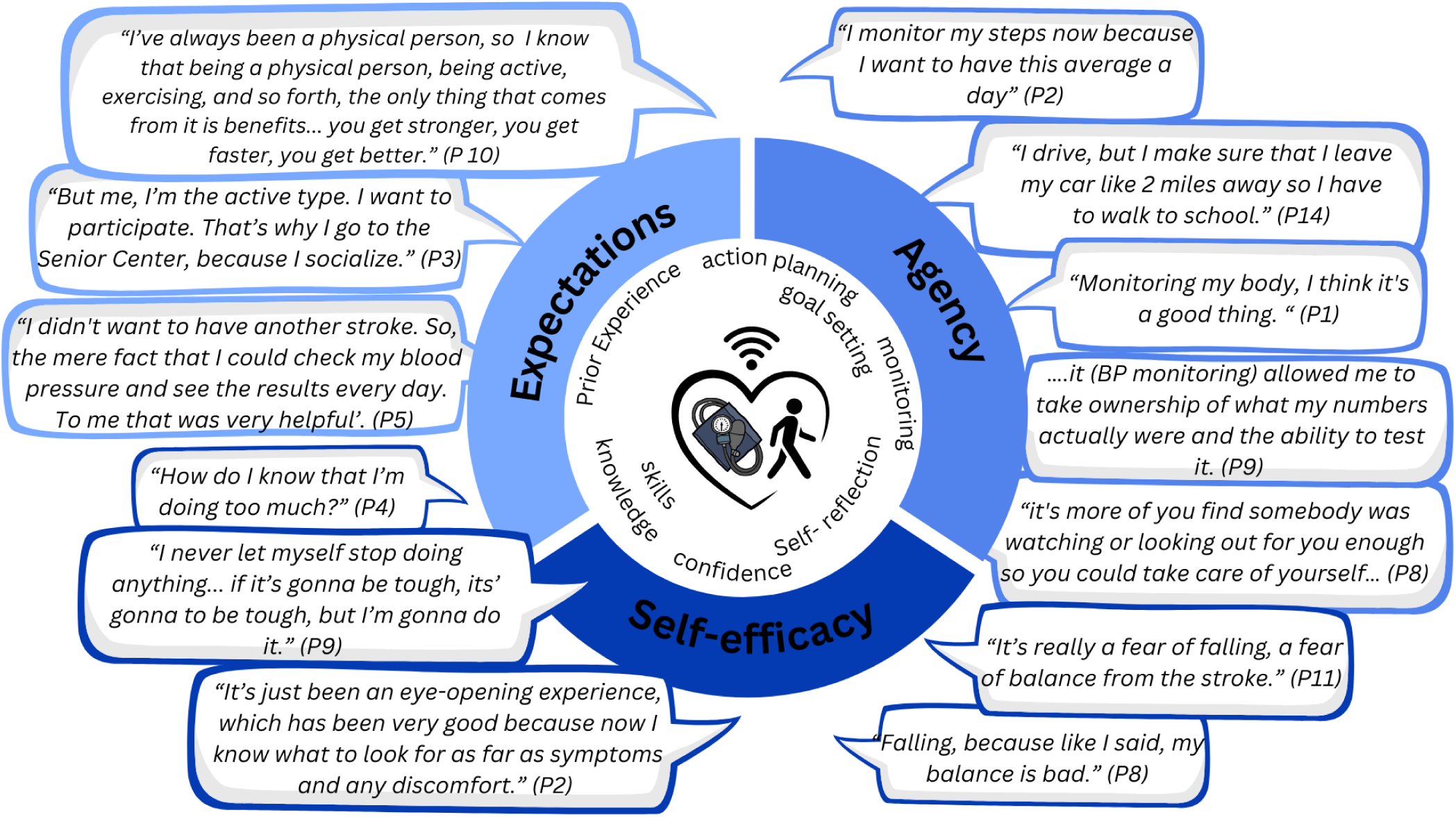
Social Cognitive Theory guided Thematic Analysis Data Display. Data displays patient quotations related to identified themes, and categories identified.

#### Data Saturation

Given the focused questions and directed interview guide, 6-12 participants is generally considered adequate to support theory-informed analysis..^42^ Saturation was tracked using an established approach^59^ that involves achieving an informational threshold of ≤5% for any new code generated that may capture characteristics for a new theme from each transcript. In line with the combined inductive-deductive approach, saturation was defined as redundancy in primary patterns rather than exhaustion of all possible meanings with the goal of reaching sufficient depth and variation of participant’s experiences. Beyond this threshold at least a few additional interviews should be conducted to ensure that saturation has been achieved. When data saturation has been achieved it signals that all themes have been identified. In our sample, data saturation was reached at 10 interviews for Interview 1 and at 6 interviews for Interview 2. (Suppl.) Additional interviews were conducted to ensure that data saturation was achieved. The scope of questions for the first interview was broad, including questions about behavior change in the TASC trial and current health behaviors and therefore required more interviews.

#### Rigor

Several strategies were used to enhance analytic rigor. These pursuits explored researcher subjectivity to enhance the rigor, or “trustworthiness” of qualitative research.^60,61^ Codes were peer-reviewed (IAN, KF) throughout the analysis and team members with diverse expertise resolved discrepancies for clarity and consistency in coding Reflexive consideration of researcher positionality, including prior experience in chronic disease prevention and work with underserved stroke populations, was documented throughout analysis to examine potential interpretive bias. Member checks were conducted to confirm contextual accuracy and resonance of findings with participants’ experiences after data analysis.^62^

The data that support the findings of this study are available from the corresponding author upon reasonable request per the journal’s Transparency and Openness Promotion Guidelines. This study was approved by CUIMC’s Institutional Review Board and reported according to Standards for Reporting Qualitative Research (SRQR) guidelines.^63^ (**Suppl**.) All participants provided written informed consent.

## RESULTS

### Participant Characteristics

Characteristics of the 14 participants (7 female, mean age 59 ± 9.6 years) are displayed in Table 1. Eight participants were Non-Hispanic Black, two were Non-Hispanic White, and four were Hispanic (3 bilingual). Of the PROs, mean (SD) PROMIS physical function T-score was 44.9 (9.8), while the mean physical function T-score for the general population is 50 (10). PA varied across IPAQ categories, with only two (14%) reporting low levels of PA. Six (43%) identified with URICA contemplation stage indicating they were planning PA behavior in the next 6 months (Table 1).

**Table 1.**
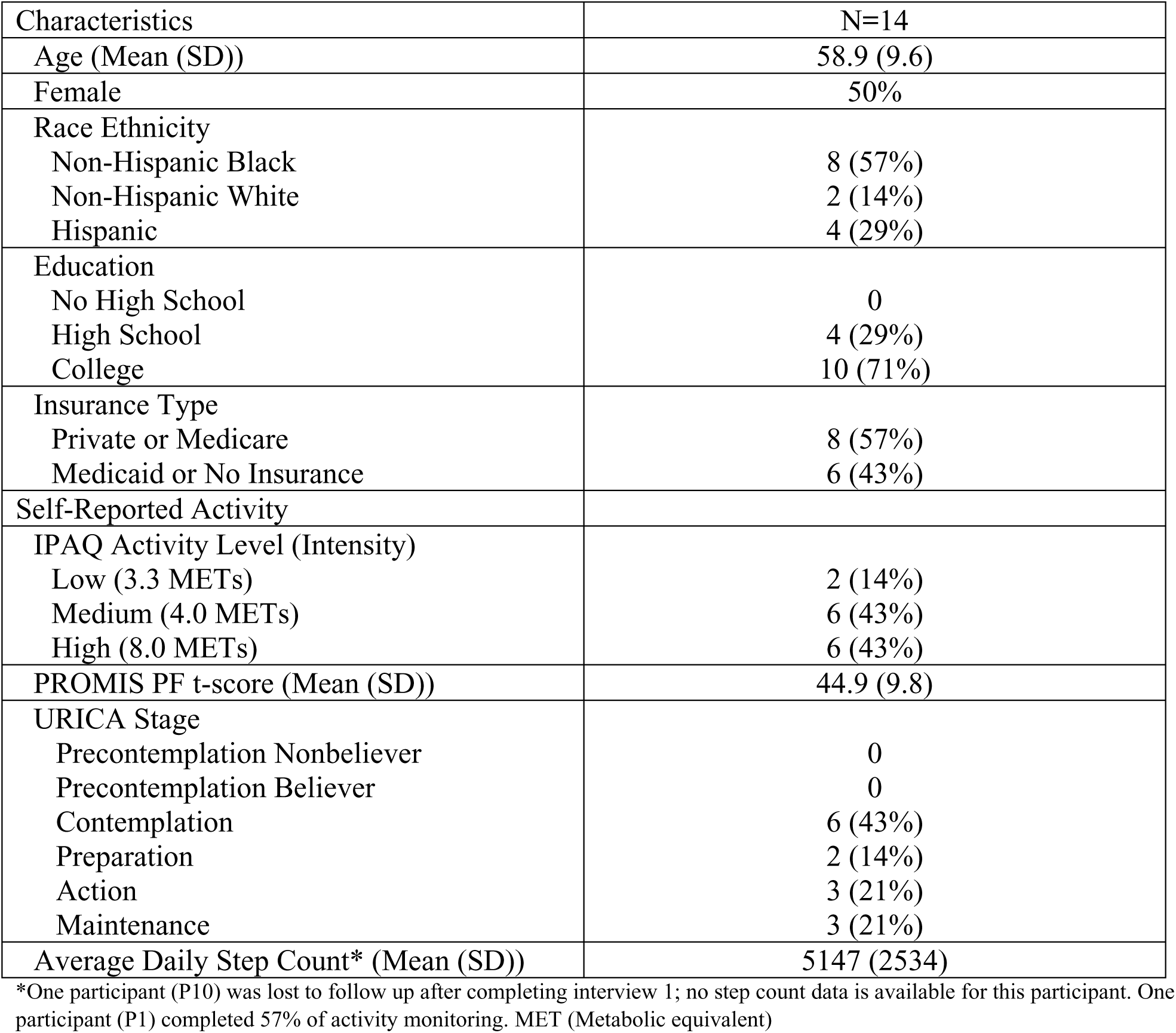
Study Participant Demographics and Patient Reported Outcomes.

|  |  |
| --- | --- |
| Characteristics | N=14 |
| Age (Mean (SD)) | 58.9 (9.6) |
| Female | 50% |
| Race Ethnicity |  |
| Non-Hispanic Black | 8 (57%) |
| Non-Hispanic White | 2 (14%) |
| Hispanic | 4 (29%) |
| Education |  |
| No High School | 0 |
| High School | 4 (29%) |
| College | 10 (71%) |
| Insurance Type |  |
| Private or Medicare | 8 (57%) |
| Medicaid or No Insurance | 6 (43%) |
| Self-Reported Activity |  |
| IPAQ Activity Level (Intensity) |  |
| Low (3.3 METs) | 2 (14%) |
| Medium (4.0 METs) | 6 (43%) |
| High (8.0 METs) | 6 (43%) |
| PROMIS PF t-score (Mean (SD)) | 44.9 (9.8) |
| URICA Stage |  |
| Precontemplation Nonbeliever | 0 |
| Precontemplation Believer | 0 |
| Contemplation | 6 (43%) |
| Preparation | 2 (14%) |
| Action | 3 (21%) |
| Maintenance | 3 (21%) |
| Average Daily Step Count* (Mean (SD)) | 5147 (2534) |

\*One participant (P10) was lost to follow up after completing interview 1; no step count data is available for this participant. One participant (P1) completed 57% of activity monitoring. MET (Metabolic equivalent)

### Blood Pressure Management

Among our participants, nine (64%) reported that their behavior changed after TASC: either self-monitoring their home BP when they never had or becoming more consistent and informed on their numbers. Three (21%) participants had monitored BP prior to the intervention and continued to do so. Only two (14%) participants, who had not previously monitored, reported no change after the intervention.

Four (29%) participants stated they were not consistently adherent with medications prior to intervention but changed habits afterward, while eleven (78%) reported medication adherence regardless of the intervention. Only one (7%) participant reported a continuous nonadherence to medication. Nine (64%) participants appreciated technological assistance when needed, with care-partner support.

### Physical Activity

In the first interview, participants reported that their primary form of PA was walking or performing household chores. Walking occurred for leisure, exercise, occupational role, or for transportation. In the second interview, all participants reported positive experience with PA monitoring. All adhered to wearing the PA monitor for the entire month, except for patient 1 who completed a little over 2 weeks in total. Nine participants (64%) required up to 2 phone calls for remote set up while 4 (29%) needed weekly calls with assistance from a care partner (sister, daughter, son, or husband) for adherence to PA monitoring. (Suppl.)

Findings from the activity monitors revealed that only three participants (21%) achieved the recommended 150 minutes of weekly MVPA, and five (36%) attained the 6000 step/day threshold protective against future cardiovascular events.^64,65^

### Qualitative Analysis

Fourteen and thirteen interviews for Interview 1 and 2, respectively, provided sufficient richness and depth for analysis. The three overarching themes were consolidated from both interviews and presented in a data display (Figure 2).

Using SCT, which highlights the multi-directional relationship among self-efficacy, outcome expectations, and socio-structural factors,^66^ we identified three key themes that influenced participation in secondary stroke prevention behaviors (Figure 3).

**Figure 3.**
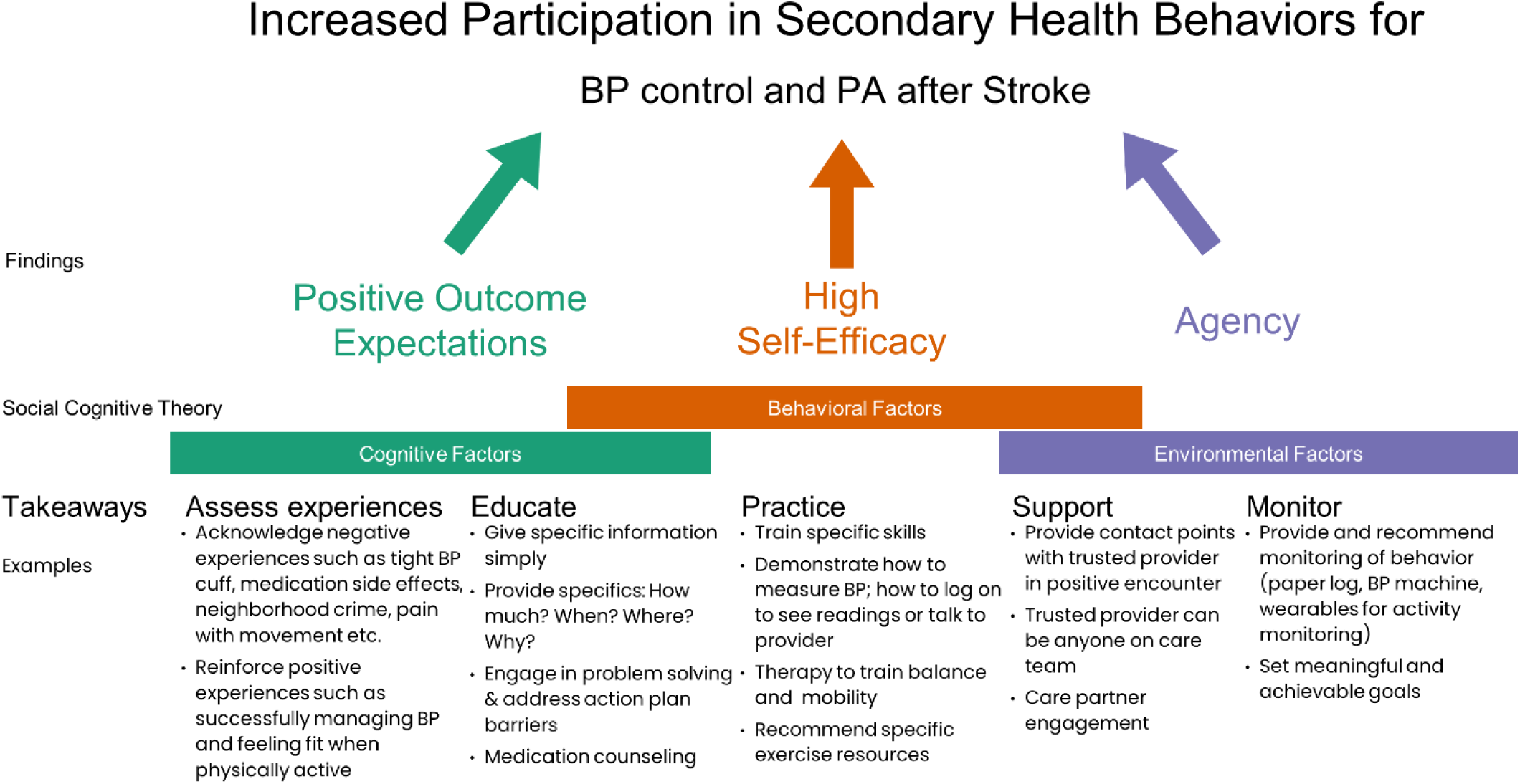
Distilled factors and takeaways to increase participation in secondary health behaviors of BP management and PA.

#### Theme 1 – Outcome Expectations

All participants believed that secondary stroke prevention behaviors would lead to positive outcomes based on their prior life experiences. Hypertension was understood to be a risk factor for stroke recurrence, one TASC participant noted, *“I have chronic high blood pressure all my life, so it was one of the factors that led to my stroke. So, me educating myself on that it really helped me to monitor it more on a more successful schedule.” (P9)* Further, BP control and medication adherence was commonly connected to reduced risk of another stroke and overall health benefits: *“I don’t want to go through that stroke thing again. So now I’m taking it [medication] every morning.” (P4)*

Experiences with physical activity revealed similar associations with stroke prevention “I have to stay active. I don’t want to suffer again what I suffered” (P6) and health-related outcomes “I’ve always been a physical person, so I know that being a physical person, being active, exercising, and so forth, the only thing that comes from it is benefits. The only thing that comes from it is you get strength, you get stronger, you get faster, you get better.” (P 10). Other expectations were more immediate, including emotional “Physical activity cheers me up” (P 11) and social well-being “And that’s why I go to the Senior Center, because I socialize.” (P3)

#### Theme 2 – Self-efficacy

Behaviors that were well-established in daily life and perceived as “mastered” persisted despite new environmental obstacles *“I used it [BP monitor]. I wasn’t uncomfortable using it because I had one already and then, so it was easy for me to adapt easily.” (P14) or* physical barriers were circumvented with purpose *“I never let myself stop doing anything. I just did it anyway. But what I did when I first went home is I used the cane. But I never let myself stop doing anything. I don’t care what I have to do. If I’m gonna do something, I’m gonna do it. If it’s gonna be tough, it’s going to be tough, but I’m gonna do it. (P9),* and *“I do a lot of walking in the neighborhood. I lift my walker and go up on the sidewalk because the walker can’t lift itself, so I lift it and go up on the sidewalk.” (P3)*

Formal, structured and specific information sharing as part of the TASC intervention fostered mechanistic understanding of the relationship between health behaviors and associated risks. “*I learned why it’s important, you know if I want to maintain good health, I have to take my medication. I have to stay on top of that. So that was the most important piece that I got from the (TASC) study*” *(P9).* For visual learners, the tailored infographic supported retention:*“I’m a visual person. If you show me something in a picture or you show me in a graphic diagram, I can understand it better then and I’ll remember more” (P8).* Consistent contact with a health professional provided a scaffold for learning new concepts “E*ach time that I talk to nurse it was more knowledge that I would learn, more information” (P1)*.

Alternatively, lack of both knowledge “*I was oblivious to certain things regarding what happens when you have high blood pressure, why do I want to maintain a low blood pressure? (P9)* and/or skills such as using a BP device *“It was good because I learned how to use it [BP monitoring]. I wasn’t doing it correctly.”* hampered participation in health behaviors. General health advice given during hospitalization such as, “*…do a lot of exercise like walking.”* (P4) was insufficient, resulting in confusion “*How do I know that I’m doing too much [physical activity]?” (P4).* An incomplete view of PA as only “structured exercise” led to low self-efficacy in the behavior *“my work [housekeeping] doesn’t allow me [time to exercise]”* (P6) who also reported being in contemplation phase for exercise, despite objectively exceeding recommended weekly MVPA and step counts.

Fear of falling was reported by over 60% of participants “*It’s really a fear falling, a fear of the balance from the stroke. (P11)* and, consequently, limited their activity *“It made me more afraid, so for some time I limited my activities.” (P10)*.

#### Theme 3 – Agency

Self-management tools were highlighted among participants as methods used to build and increase adherence to positive health behaviors. Individuals mentioned stacking habits. BP monitoring promoted medication adherence “*So now I’m taking it [blood pressure] every morning. I just have it with my other medication together. So that’s the first thing” (P4) “But now since I’m in a routine and I have a regimen of when to do it or when to take my stuff, my medication and my blood pressure it’s not a problem. I was thinking what else can I do?.. it reminds me every morning about my meditation.” (P2)* and building behaviors into their daily life even when less challenging options were available *“Instead of taking the [subway]train, I walk.” (P6) “I drive but I make sure that I leave my car like 2 miles away so that I have to walk to school.” (P14)*

Intentional forethought and action planning were used to circumvent personal barriers *“I have a little case that I put all my medication in by day. Just go to that case and take those pills out and take them. I don’t have to be looking for them. Because they are organized already. I just go put them in my hand and take them.” (P1),* and socio-structural obstacles such as stairs, public transportation and neighborhood crime *“Unfortunately, we have a lot of crime over here, so I have to time it [PA] and everything; like basically mid-morning when everybody’s out and about.’ (P9)*

Self-monitoring routinized as a direct consequence of all active telemonitoring behaviors, *“To this day I have a blood pressure machine, I have a glucometer, I have the oxygen thing for my finger. So, I keep track of all my numbers….it allowed me to take ownership of what my numbers actually were and the ability to test it.” (P9*) and *“I monitor my steps now because I want to have this average [number of steps] a day.” (P2)*. Consciously setting goals with the intent to maintain successful behavior was integral to creating the perception of self-control: *“kind of monitor myself like (it promotes) self-discipline.” (P7) and “It gives you an idea of what you need to do daily for yourself”.” (P8)*

Among our participants, social support provided after acute stroke complemented and supported personal agency. *“The providers were not the solution; it’s doing what the providers asked me to do…enhancing motivation and not feeling sorry for myself” (P14).* Encouragement from care-partners was contributory *“more recommendation from my oldest daughter saying that they needed something to monitor me.” (P2)* However, when participants felt that their autonomy was limited by care-partners the likelihood of performing positive health behaviors diminished: “*My wife. She don’t want me to go no place unless she was there with me. She didn’t want me to go no place by myself.” (P1); “I feel like I’m on lockdown. That’s the barrier. I can’t go anywhere unaccompanied.” (P3)*.

## DISCUSSION

Participation in multiple behaviors including BP management and PA appeared to be predominantly influenced by individual factors of outcome expectations, self-efficacy, and agency. The TASC intervention with tailored BP infographics followed by the practice of BP monitoring with devices for self-management allowed individuals to reflect on their own risk, current behavior, and begin to strategize a path forward with acquired knowledge and attained skills. It served to reconceptualize these healthy behaviors and allowed participants to establish ways to integrate them into daily life, i.e. build multiple healthy habits.^67,68^ Therefore, patient-centered behavior change intervention design should consider contextualizing these factors for best clinical outcomes.

### Positive outcome expectations

Outcome expectations are the anticipated consequences of engaging in a particular behavior. However, belief in positive outcomes is insufficient to create and sustain change on its own.^69^ Our TASC participants aware of how their acquired knowledge tied with positive outcomes of secondary stroke prevention developed BP self-management behaviors to bridge the gap between awareness and action, i.e. cognizant pursuit of positive outcome expectations.

PROs with objective data allowed us to contextualize patient perceptions of PA to their performance in the community. Although our participants were cognizant of the importance of PA, they did not have adequate knowledge to recognize or implement the behavior successfully. Our findings substantiate evidence that general education for rehabilitation provided in the hospital is not sufficient to promote PA behaviors.^70–72^

### Self-efficacy

Self-efficacy is specifically tied to belief in one’s ability to successfully commit to a behavior despite barriers. It is the critical behavioral factor that interacts reciprocally between personal and environmental factors to modulate health behaviors.^73^ Higher self-efficacy is known to have a positive correlation with a higher level of treatment adherence.^74^ Our TASC participants built a high sense of self-efficacy in their own BP management with medication adherence as they successfully monitored their BP with a device, prompting positive outcomes of good BP control.

Those with higher self-efficacy for PA behaviors were more likely to see socio-structural barriers as steppingstones instead of hurdles. However, environmental factors alone do not fully explain PA participation.^75–77^ A majority also reported low balance self-efficacy and fear of falling. Regardless of actual physical disability, self-efficacy predicts who can participate in and achieve the greatest gains in mobility.^78–81^ Such fear can limit PA post stroke.^82,83^

### Agency

We learned that monitoring is a critical component of behavior change,^84^ but it is not sufficient in isolation.^22^ Monitoring tools were well received, as they provided a tangible means to self-evaluate, set intentions, and establish goals. Agency or perceived control was developed not only through consistent use of these tools but also meaningful contact with trusted health professionals. This supported environment provided safe space to foster agency for active problem solving and self-reflection, allowing individuals to evaluate their current behavior, assess their ability to reach goals and set their intentions to align with recommendations. As noted in SCT, self-reflection allows for evaluation and corrective readjustment to meet goals.^85^

Social support and personal agency may be seen as a substitute for one another or a complement to one another in the context of health behaviors.^86^ For example, in the former, where support is a substitute for agency, a person who knows that a health professional will alert them if there is an issue with their blood pressure, might take less ownership in monitoring themselves and learning critical BP responses.

Care-partners also influenced stroke survivor agency. These care partners may carry their own emotional burden and gaps in knowledge limiting their positive outcomes expectations and agency.^87^ Therefore, care-partners should be engaged early with health professional teams during transitions of care to ensure complementary and enhanced agency.

### Proposed Behavioral Mechanism and Clinical Implications

In contrast to passive information sharing, priming behavior changes with actively engaged education and directed problem solving is vital to promote positive outcome expectations and advance specific skills for optimal self-efficacy.^66,88,89^ Further, agency or perceived control as telemonitored feedback may sustain behavior change.

Once agency is established, multiple behavior changes may be possible in the post-acute stroke period for a successful behavioral intervention.^90–93^ Multiple healthy habit formation can be encouraged simultaneously or developed sequentially.^94^ Forming habits such as checking BP together with taking medication, i.e. habit stacking, can improve consistency by adding meaning to the task, and decrease the attention (cognitive load) required to initiate the task. This is the strength of habitual automaticity with cue-triggered responses driving behavior with little intentional effort.^95,96^ A cue to action such as the ‘teachable moment’ instance through manifestation of the perceived threat from stroke, has the potential to trigger healthy behaviors through clinician-patient interaction.^97^ Reframing reward expectations to increase subjective value of larger, distant, and patient-oriented outcomes such as reduced risk of recurrent stroke and better recovery may reduce cognitive burden to routinize these health behavior tasks.^98^ Thus, we hypothesized a conceptual model with mechanistic targets to optimize multimodal health behavior uptake after stroke for telehealth-based behavioral interventions (Figure S1).

Long term support may be harnessed through coordinated health services to include care partners. Community stroke support groups^99^ have been encouraged by prominent stroke associations to support recovery.^100^ These can serve to provide multidisciplinary knowledge and skills training, to raise awareness among patients/care partners regarding local resources, and to leverage those chronic stroke patients with sustained improvements in health behaviors to support their early post-stroke peers. Key takeaways with suggested examples noted in Figure 3.

## Limitations

This study has several limitations. We acknowledge that our small sample size limits generalizability. However, with our strict inclusion criteria, this was adequate for a qualitative study with semi-structured interviews that we continued to conduct interviews even after data saturation was reached. The interviewer as a research instrument being a physician may also have limited discussions. However, to counter this bias research subjectivity was explored as part of qualitative data validity and reliability measures. For a Spanish only speaking participant, another Spanish speaking interviewer was utilized, and coding was conducted on the translated script. This could have restricted the data retrieved. Patient interviews may also have been limited by recall bias and time elapsed since the event. However, most participants had a clear recollection of their event, the impact of TASC and time course along the stroke continuum of care. Although the recruited participants may have been those willing to engage, lessons gleaned from these individuals provided an understanding of components most successful for behavior change. Further, our results are consistent with previous literature on behavior change. This provides reassurance that our findings are generalizable to mild stroke survivors in an underserved setting.

## CONCLUSION

We learned that consistent participation in secondary stroke prevention behaviors is more likely when positive outcome expectations, self-efficacy, and agency are high. Further, BP and PA monitoring with feedback improved agency with complementary social support. Targeted education to drive knowledge and skill development with trusted support, and monitoring tools may be integral components to initiate lasting preventive behaviors after stroke.

## Data Availability

Data will be made available from the corresponding author upon reasonable request.

## Acknowledgements

The authors acknowledge study coordinator Carmen Castillo, all the participants and their care partners who came together to make this study possible.

## Nonstandard Abbreviations and Acronyms

BP: Blood Pressure
PA: Physical Activity
TASC: Telehealth After Stroke Care

## Supplemental Material

Reporting Checklist

Methods and Materials S1-S3

Figure S1-2

Table S1

## REFERENCES

1. Kleindorfer DO, Towfighi A, Chaturvedi S, Cockroft KM, Gutierrez J, Lombardi-Hill D, Kamel H, Kernan WN, Kittner SJ, Leira EC, et al. 2021 Guideline for the Prevention of Stroke in Patients With Stroke and Transient Ischemic Attack: A Guideline From the American Heart Association/American Stroke Association. Stroke. 2021;52. doi: 10.1161/STR.0000000000000375

2. Albright KC, Huang L, Blackburn J, Howard G, Mullen M, Bittner V, Muntner P, Howard V. Racial differences in recurrent ischemic stroke risk and recurrent stroke case fatality. Neurology. 2018;91:e1741–e1750. doi: 10.1212/WNL.0000000000006467

3. Park J-H, Ovbiagele B. Association of black race with recurrent stroke risk. Journal of the Neurological Sciences. 2016;365:203–206. doi: 10.1016/j.jns.2016.04.012

4. Samsa GP, Bian J, Lipscomb J, Matchar DB. Epidemiology of Recurrent Cerebral Infarction: A Medicare Claims–Based Comparison of First and Recurrent Strokes on 2-Year Survival and Cost. Stroke. 1999;30:338–349. doi: 10.1161/01.STR.30.2.338

5. Hackam DG, Spence JD. Combining multiple approaches for the secondary prevention of vascular events after stroke: a quantitative modeling study. Stroke. 2007;38:1881–1885. doi: 10.1161/strokeaha.106.475525

6. Mozaffarian D, Benjamin EJ, Go AS, Arnett DK, Blaha MJ, Cushman M, de Ferranti S, Despres JP, Fullerton HJ, Howard VJ, et al. Heart disease and stroke statistics-2015 update: a report from the american heart association. Circulation. 2015;131:e29–e322. doi: 10.1161/CIR.0000000000000152

7. Brenner DA, Zweifler RM, Gomez CR, Kissela BM, Levine D, Howard G, Coull B, Howard VJ. Awareness, treatment, and control of vascular risk factors among stroke survivors. J Stroke Cerebrovasc Dis. 2010;19:311–320. doi: 10.1016/j.jstrokecerebrovasdis.2009.07.001

8. White CL, Pergola PE, Szychowski JM, Talbert R, Cervantes-Arriaga A, Clark HD, Del Brutto OH, Godoy IE, Hill MD, Pelegri A, et al. Blood pressure after recent stroke: baseline findings from the secondary prevention of small subcortical strokes trial. Am J Hypertens. 2013;26:1114–1122. doi: 10.1093/ajh/hpt076

9. Siegel D. Barriers to and strategies for effective blood pressure control. Vascular health and risk management. 2005;1:9–14. doi: 10.2147/vhrm.1.1.9.58940

10. Ireland SE, Arthur HM, Gunn EA, Oczkowski W. Stroke prevention care delivery: predictors of risk factor management outcomes. Int J Nurs Stud. 2011;48:156–164. doi: 10.1016/j.ijnurstu.2010.07.003

11. Aronow WS, Fleg JL, Pepine CJ, Artinian NT, Bakris G, Brown AS, Ferdinand KC, Ann Forciea M, Frishman WH, Jaigobin C, et al. ACCF/AHA 2011 expert consensus document on hypertension in the elderly: a report of the American College of Cardiology Foundation Task Force on Clinical Expert Consensus Documents developed in collaboration with the American Academy of Neurology, American Geriatrics Society, American Society for Preventive Cardiology, American Society of Hypertension, American Society of Nephrology, Association of Black Cardiologists, and European Society of Hypertension. Journal of the American Society of Hypertension : JASH. 2011;5:259–352. doi: 10.1016/j.jash.2011.06.001

12. White CL, Pergola PE, Szychowski JM, Talbert R, Cervantes-Arriaga A, Clark HD, Del Brutto OH, Godoy IE, Hill MD, Pelegrí A, et al. Blood pressure after recent stroke: baseline findings from the secondary prevention of small subcortical strokes trial. Am J Hypertens. 2013;26:1114–1122. doi: 10.1093/ajh/hpt076

13. Sardana M, Lin H, Zhang Y, Liu C, Trinquart L, Benjamin EJ, Manders ES, Fusco K, Kornej J, Hammond MM, et al. Association of Habitual Physical Activity With Home Blood Pressure in the Electronic Framingham Heart Study (eFHS): Cross-sectional Study. J Med Internet Res. 2021;23:e25591. doi: 10.2196/25591

14. Billinger SA, Arena R, Bernhardt J, Eng JJ, Franklin BA, Johnson CM, MacKay-Lyons M, Macko RF, Mead GE, Roth EJ, et al. Physical Activity and Exercise Recommendations for Stroke Survivors. Stroke. 2014;45:2532–2553. doi: doi:10.1161/STR.0000000000000022

15. Fini NA, Holland AE, Keating J, Simek J, Bernhardt J. How Physically Active Are People Following Stroke? Systematic Review and Quantitative Synthesis. Phys Ther. 2017;97:707–717. doi: 10.1093/ptj/pzx038

16. English C, Manns PJ, Tucak C, Bernhardt J. Physical Activity and Sedentary Behaviors in People With Stroke Living in the Community: A Systematic Review. Physical Therapy. 2014;94:185–196. doi: 10.2522/ptj.20130175

17. Grady PA, Gough LL. Self-Management: A Comprehensive Approach to Management of Chronic Conditions. American Journal of Public Health. 2014;104.

18. Sakakibara BM, Kim AJ, Eng JJ. A Systematic Review and Meta-Analysis on Self-Management for Improving Risk Factor Control in Stroke Patients. International journal of behavioral medicine. 2017;24:42–53. doi: 10.1007/s12529-016-9582-7

19. Boysen G, Krarup L-H, Zeng X, Oskedra A, Korv J, Andersen G, Gluud C, Pedersen A, Lindahl M, Hansen L, et al. ExStroke Pilot Trial of the effect of repeated instructions to improve physical activity after ischaemic stroke: a multinational randomised controlled clinical trial. BMJ. 2009;339:b2810–b2810. doi: 10.1136/bmj.b2810

20. Cheng D, Qu Z, Huang J, Xiao Y, Luo H, Wang J. Motivational interviewing for improving recovery after stroke. Cochrane Database Syst Rev. 2015;2015:Cd011398. doi: 10.1002/14651858.CD011398.pub2

21. Marzolini S. Clinician’s Commentary on Hui et al.(1). Physiother Can. 2018;70:90–91. doi: 10.3138/ptc.2016-40-cc

22. Lynch EA, Jones TM, Simpson DB, Fini NA, Kuys SS, Borschmann K, Kramer S, Johnson L, Callisaya ML, Mahendran N, et al. Activity monitors for increasing physical activity in adult stroke survivors. Cochrane Database Syst Rev. 2018;7:Cd012543. doi: 10.1002/14651858.CD012543.pub2

23. Cao C, Jain N, Lu E, Sajatovic M, Still CH. Secondary Stroke Risk Reduction in Black Adults: a Systematic Review. Journal of Racial and Ethnic Health Disparities. 2023;10:306–318. doi: 10.1007/s40615-021-01221-2

24. McKenna S, Jones F, Glenfield P, Lennon S. Bridges self-management program for people with stroke in the community: A feasibility randomized controlled trial. Int J Stroke. 2015;10:697–704. doi: 10.1111/ijs.12195

25. Hall P, Lawrence M, Blake C, Lennon O. Interventions for Behaviour Change and Self-Management of Risk in Stroke Secondary Prevention: An Overview of Reviews. Cerebrovasc Dis. 2024;53:1–13. doi: 10.1159/000531138

26. Reeves MJ, Hughes AK, Woodward AT, Freddolino PP, Coursaris CK, Swierenga SJ, Schwamm LH, Fritz MC. Improving transitions in acute stroke patients discharged to home: the Michigan stroke transitions trial (MISTT) protocol. BMC Neurol. 2017;17:115. doi: 10.1186/s12883-017-0895-1

27. Dong C, Gardener H, Rundek T, Marulanda E, Gutierrez CM, Campo-Bustillo I, Gordon Perue G, Johnson KH, Sacco RL, Romano JG. Factors and Behaviors Related to Successful Transition of Care After Hospitalization for Ischemic Stroke. Stroke. 2023;54:468–475. doi: 10.1161/strokeaha.122.040891

28. Naqvi IA, Strobino K, Cheung YK, Li H, Schmitt K, Ferrara S, Tom SE, Arcia A, Williams OA, Kronish IM, et al. Telehealth After Stroke Care Pilot Randomized Trial of Home Blood Pressure Telemonitoring in an Underserved Setting. Stroke. 2022;53:3538–3547. doi: doi:10.1161/STROKEAHA.122.041020

29. Reeves MJ, Boden-Albala B, Cadilhac DA. Care Transition Interventions to Improve Stroke Outcomes: Evidence Gaps in Underserved and Minority Populations. Stroke. 2023;54:386–395. doi: doi:10.1161/STROKEAHA.122.039565

30. Shippee ND, Shah ND, May CR, Mair FS, Montori VM. Cumulative complexity: a functional, patient-centered model of patient complexity can improve research and practice. J Clin Epidemiol. 2012;65:1041–1051. doi: 10.1016/j.jclinepi.2012.05.005

31. Wood JP, Connelly DM, Maly MR. ’Getting back to real living’: A qualitative study of the process of community reintegration after stroke. Clin Rehabil. 2010;24:1045–1056. doi: 10.1177/0269215510375901

32. Naqvi IA, Cheung YK, Strobino K, Li H, Tom SE, Husaini Z, Williams OA, Marshall RS, Arcia A, Kronish IM, et al. TASC (Telehealth After Stroke Care): a study protocol for a randomized controlled feasibility trial of telehealth-enabled multidisciplinary stroke care in an underserved urban setting. Pilot Feasibility Stud. 2022;8:81. doi: 10.1186/s40814-022-01025-z

33. Naqvi IA SK, Li H, Schmitt K, Barratt Y, Ferrara S, Hasni A, Cato KD, Weiner MG, Elkind MSV, Kronish IM, Arcia A. Improving Patient-Reported Outcomes in Stroke Care using Remote Blood Pressure Monitoring and Telehealth. Appl Clin Inform. 2023;doi: 10.1055/s-0043-1772679. doi: 10.1055/s-0043-1772679.

34. Michie S, Johnston M, Francis J, Hardeman W, Eccles M. From theory to intervention: Mapping theoretically derived behavioural determinants to behaviour change techniques. Applied Psychology: An International Review. 2008;57:660–680. doi: 10.1111/j.1464-0597.2008.00341.x

35. da Cruz Peniche P, Faria C, Hall P, Lennon O. Effectiveness of behavior change and self-management theoretically-informed telehealth interventions for stroke secondary prevention: An overview of systematic reviews. J Telemed Telecare. 2024:1357633x241238779. doi: 10.1177/1357633x241238779

36. Salinas J, Schwamm LH. Behavioral Interventions for Stroke Prevention. Stroke. 2017;48:1706–1714. doi: doi:10.1161/STROKEAHA.117.015909

37. Bandura A. Social foundations of thought and action: A social cognitive theory. Englewood Cliffs, NJ, US: Prentice-Hall, Inc; 1986.

38. Li Y, Feng X, Zhang M, Zhou M, Wang N, Wang L. Clustering of cardiovascular behavioral risk factors and blood pressure among people diagnosed with hypertension: a nationally representative survey in China. Scientific Reports. 2016;6:27627. doi: 10.1038/srep27627

39. Schuit AJ, Van Loon AJM, Tijhuis M, Ocké MC. Clustering of Lifestyle Risk Factors in a General Adult Population. Preventive Medicine. 2002;35:219–224. doi: 10.1006/pmed.2002.1064

40. Etikan I, Musa SA, Alkassim RS. Comparison of Convenience Sampling and Purposive Sampling. American Journal of Theoretical and Applied Statistics. 2016;5:1.

41. Kuper A, Lingard L, Levinson W. Critically appraising qualitative research. Bmj. 2008;337:a1035. doi: 10.1136/bmj.a1035

42. Sandelowski M. Sample size in qualitative research. Research in Nursing & Health. 1995;18:179–183. doi: 10.1002/nur.4770180211

43. Naqvi IA, Strobino K, Kuen Cheung Y, Li H, Schmitt K, Ferrara S, Tom SE, Arcia A, Williams OA, Kronish IM, et al. Telehealth After Stroke Care Pilot Randomized Trial of Home Blood Pressure Telemonitoring in an Underserved Setting. Stroke. 2022;53:3538–3547. doi: 10.1161/STROKEAHA.122.041020

44. Support Z. HIPAA Business Associate Agreement (BAA). https://support.zoom.com/hc/en/article?id=zm_kb&sysparm_article=KB0067751. 2025. Accessed 1/15/2025.

45. Microsoft. Microsoft Stream. https://support.microsoft.com/en-us/office/learn-more-about-stream-on-sharepoint-cf4c10c8-5ed3-4229-9e2a-60d31b31575d. 2025. Accessed 1/15/25.

46. Katzan I, Schuster A, Kinzy T. Physical Activity Monitoring Using a Fitbit Device in Ischemic Stroke Patients: Prospective Cohort Feasibility Study. JMIR Mhealth Uhealth. 2021;9:e14494. doi: 10.2196/14494

47. Fulk GD, Combs SA, Danks KA, Nirider CD, Raja B, Reisman DS. Accuracy of 2 activity monitors in detecting steps in people with stroke and traumatic brain injury. Phys Ther. 2014;94:222–229. doi: 10.2522/ptj.20120525

48. Hui J, Heyden R, Bao T, Accettone N, McBay C, Richardson J, Tang A. Validity of the Fitbit One for Measuring Activity in Community-Dwelling Stroke Survivors. Physiother Can. 2018;70:81–89. doi: 10.3138/ptc.2016-40.ep

49. Katzan IL, Fan Y, Uchino K, Griffith SD. The PROMIS physical function scale. A promising scale for use in patients with ischemic stroke. 2016;86:1801–1807. doi: 10.1212/wnl.0000000000002652

50. HealthMeasures Scoring Service powered by Assessment Center. An application to score PROMIS®, NIH Toolbox®, and Neuro-QoL™ instruments. https://www.assessmentcenter.net/ac_scoringservice.

51. Fini NA, Simpson D, Moore SA, Mahendran N, Eng JJ, Borschmann K, Moulaee Conradsson D, Chastin S, Churilov L, English C. How should we measure physical activity after stroke? An international consensus. Int J Stroke. 2023;18:1132–1142. doi: 10.1177/17474930231184108

52. Craig CL, Marshall AL, Sjöström M, Bauman AE, Booth ML, Ainsworth BE, Pratt M, Ekelund U, Yngve A, Sallis JF, et al. International physical activity questionnaire: 12-country reliability and validity. Med Sci Sports Exerc. 2003;35:1381–1395. doi: 10.1249/01.Mss.0000078924.61453.Fb

53. Lerdal A, Moe B, Digre E, Harding T, Kristensen F, Grov EK, Bakken LN, Eklund ML, Ruud I, Rossi JS. Stages of Change--continuous measure (URICA-E2): psychometrics of a Norwegian version. J Adv Nurs. 2009;65:193–202. doi: 10.1111/j.1365-2648.2008.04842.x

54. Fitabase. https://www.fitabase.com/. Accessed 12/05/2024.

55. Orstad SL, Gerchow L, Patel NR, Reddy M, Hernandez C, Wilson DK, Jay M. Defining Valid Activity Monitor Data: A Multimethod Analysis of Weight-Loss Intervention Participants’ Barriers to Wear and First 100 Days of Physical Activity. Informatics. 2021;8:39. doi: 10.3390/informatics8020039

56. Chan A, Chan D, Lee H, Ng CC, Yeo AHL. Reporting adherence, validity and physical activity measures of wearable activity trackers in medical research: A systematic review. Int J Med Inform. 2022;160:104696. doi: 10.1016/j.ijmedinf.2022.104696

57. Semanik P, Lee J, Pellegrini CA, Song J, Dunlop DD, Chang RW. Comparison of Physical Activity Measures Derived From the Fitbit Flex and the ActiGraph GT3X+ in an Employee Population With Chronic Knee Symptoms. ACR Open Rheumatology. 2020;2:48–52. doi: 10.1002/acr2.11099

58. 9.2.22 DV. Cloud application for managing, analyzing, and presenting qualitative and mixed method research data SocioCultural Research Consultants, LLC. www.dedoose.com. 2024. Accessed 12/05/2024.

59. Guest G, Namey E, Chen M. A simple method to assess and report thematic saturation in qualitative research. PLoS One. 2020;15:e0232076. doi: 10.1371/journal.pone.0232076

60. Bradbury-Jones C. Enhancing rigour in qualitative health research: exploring subjectivity through Peshkin’s I’s. J Adv Nurs. 2007;59:290–298. doi: 10.1111/j.1365-2648.2007.04306.x

61. Guba EG. Criteria for assessing the trustworthiness of naturalistic inquiries. ECTJ. 1981;29:75–91. doi: 10.1007/BF02766777

62. Lincoln YS, Guba EG, Pilotta JJ. Naturalistic inquiry: Beverly Hills, CA: Sage Publications, 1985, 416 pp., $25.00 (Cloth). International Journal of Intercultural Relations. 1985;9:438–439. doi: 10.1016/0147-1767(85)90062-8

63. O’Brien BC, Harris IB, Beckman TJ, Reed DA, Cook DA. Standards for reporting qualitative research: a synthesis of recommendations. Acad Med. 2014;89:1245–1251. doi: 10.1097/acm.0000000000000388

64. Paluch AE, Bajpai S, Ballin M, Bassett DR, Buford TW, Carnethon MR, Chernofsky A, Dooley EE, Ekelund U, Evenson KR, et al. Prospective Association of Daily Steps With Cardiovascular Disease: A Harmonized Meta-Analysis. Circulation. 2023;147:122–131. doi: doi:10.1161/CIRCULATIONAHA.122.061288

65. Kono Y, Kawajiri H, Kamisaka K, Kamiya K, Akao K, Asai C, Inuzuka K, Yamada S. Predictive impact of daily physical activity on new vascular events in patients with mild ischemic stroke. Int J Stroke. 2015;10:219–223. doi: 10.1111/ijs.12392

66. Beauchamp MR, Crawford KL, Jackson B. Social cognitive theory and physical activity: Mechanisms of behavior change, critique, and legacy. Psychology of Sport and Exercise. 2019;42:110–117. doi: 10.1016/j.psychsport.2018.11.009

67. Bailey RR, Stevenson JL. How Adults With Stroke Conceptualize Physical Activity: An Exploratory Qualitative Study. The American Journal of Occupational Therapy. 2021;75:7502345010. doi: 10.5014/ajot.2021.041780

68. Wood W, Neal DT. Healthy through habit: Interventions for initiating & maintaining health behavior change. Behavioral Science & Policy. 2016;2:71 – 83.

69. Michaelsen MM, Esch T. Understanding health behavior change by motivation and reward mechanisms: a review of the literature. Front Behav Neurosci. 2023;17:1151918. doi: 10.3389/fnbeh.2023.1151918

70. Arlinghaus KR, Johnston CA. Advocating for Behavior Change With Education. Am J Lifestyle Med. 2018;12:113–116. doi: 10.1177/1559827617745479

71. Bridgwood B, Lager KE, Mistri AK, Khunti K, Wilson AD, Modi P. Interventions for improving modifiable risk factor control in the secondary prevention of stroke. Cochrane Database Syst Rev. 2018;5:Cd009103. doi: 10.1002/14651858.CD009103.pub3

72. Hendrickx W, Vlietstra L, Valkenet K, Wondergem R, Veenhof C, English C, Pisters MF. General lifestyle interventions on their own seem insufficient to improve the level of physical activity after stroke or TIA: a systematic review. BMC Neurol. 2020;20:168. doi: 10.1186/s12883-020-01730-3

73. Bandura A. Social foundations of thought and action: A social cognitive theory. Englewood Cliffs, NJ, US: Prentice-Hall, Inc; 1986.

74. Kara S. General self-efficacy and hypertension treatment adherence in Algerian private clinical settings. J Public Health Afr. 2022;13:2121. doi: 10.4081/jphia.2022.2121

75. Durcan S, Flavin E, Horgan F. Factors associated with community ambulation in chronic stroke. Disability and rehabilitation. 2016;38:245–249. doi: 10.3109/09638288.2015.1035460

76. Miller A, Pohlig RT, Reisman DS. Relationships Among Environmental Variables, Physical Capacity, Balance Self-Efficacy, and Real-World Walking Activity Post-Stroke. Neurorehabilitation and Neural Repair. 2022;36:535–544. doi: 10.1177/15459683221115409

77. Schmid AA, Van Puymbroeck M, Altenburger PA, Dierks TA, Miller KK, Damush TM, Williams LS. Balance and Balance Self-Efficacy Are Associated With Activity and Participation After Stroke: A Cross-Sectional Study in People With Chronic Stroke. Archives of Physical Medicine and Rehabilitation. 2012;93:1101–1107. doi: 10.1016/j.apmr.2012.01.020

78. Miller A, Pohlig RT, Reisman DS. Relationships Among Environmental Variables, Physical Capacity, Balance Self-Efficacy, and Real-World Walking Activity Post-Stroke. Neurorehabil Neural Repair. 2022;36:535–544. doi: 10.1177/15459683221115409

79. Morris JH, Oliver T, Kroll T, Joice S, Williams B. Physical activity participation in community dwelling stroke survivors: synergy and dissonance between motivation and capability. A qualitative study. Physiotherapy. 2017;103:311–321. doi: 10.1016/j.physio.2016.05.001

80. Espernberger KR, Fini NA, Peiris CL. Personal and social factors that influence physical activity levels in community-dwelling stroke survivors: A systematic review of qualitative literature. Clinical Rehabilitation. 2021;35:1044–1055. doi: 10.1177/0269215521993690

81. French MA, Moore MF, Pohlig R, Reisman D. Self-efficacy Mediates the Relationship between Balance/Walking Performance, Activity, and Participation after Stroke. Topics in Stroke Rehabilitation. 2016;23:77–83. doi: 10.1080/10749357.2015.1110306

82. Gazibara T, Kurtagic I, Kisic-Tepavcevic D, Nurkovic S, Kovacevic N, Gazibara T, Pekmezovic T. Falls, risk factors and fear of falling among persons older than 65 years of age: Falls in the older population. Psychogeriatrics. 2017;17:215–223. doi: 10.1111/psyg.12217

83. Landers MR, Oscar S, Sasaoka J, Vaughn K. Balance Confidence and Fear of Falling Avoidance Behavior Are Most Predictive of Falling in Older Adults: Prospective Analysis. Physical Therapy. 2016;96:433–442. doi: 10.2522/ptj.20150184

84. Page EJ, Massey AS, Prado-Romero PN, Albadawi S. The Use of Self-Monitoring and Technology to Increase Physical Activity: A Review of the Literature. Perspect Behav Sci. 2020;43:501–514. doi: 10.1007/s40614-020-00260-0

85. Bandura A. Human Agency in Social Cognitive Theory. American Psychologist. 1989.

86. Fishbach A. Personal Agency and Social Support: Substitutes of Complements? Psychological Inquiry. 2022;33:42–45. doi: 10.1080/1047840X.2022.2037999

87. Cameron TM, Walker MF, Fisher RJ. A Qualitative Study Exploring the Lives and Caring Practices of Young Carers of Stroke Survivors. International journal of environmental research and public health. 2022;19. doi: 10.3390/ijerph19073941

88. Crocker TF, Brown L, Lam N, Wray F, Knapp P, Forster A. Information provision for stroke survivors and their carers. Cochrane Database Syst Rev. 2021;11:Cd001919. doi: 10.1002/14651858.CD001919.pub4

89. Sakakibara BM, Kim AJ, Eng JJ. A Systematic Review and Meta-Analysis on Self-Management for Improving Risk Factor Control in Stroke Patients. International journal of behavioral medicine. 2017;24:42–53. doi: 10.1007/s12529-016-9582-7

90. Glanz K, Bishop DB. The Role of Behavioral Science Theory in Development and Implementation of Public Health Interventions. Annual Review of Public Health. 2010;31:399–418. doi: 10.1146/annurev.publhealth.012809.103604

91. Bandura A. Self-efficacy: The exercise of control. New York, NY, US: W H Freeman/Times Books/ Henry Holt & Co; 1997.

92. Wood W, Neal DT. A new look at habits and the habit-goal interface. Psychological Review. 2007;114:843 – 863.

93. Radu PT, Yi R, Bickel WK, Gross JJ, McClure SM. A mechanism for reducing delay discounting by altering temporal attention. J Exp Anal Behav. 2011;96:363–385. doi: 10.1901/jeab.2011.96-363

94. Hyman DJ, Pavlik VN, Taylor WC, Goodrick GK, Moye L. Simultaneous vs Sequential Counseling for Multiple Behavior Change. Archives of Internal Medicine. 2007;167:1152–1158. doi: 10.1001/archinte.167.11.1152

95. Orbell S, Verplanken B. The automatic component of habit in health behavior: Habit as cue-contingent automaticity. Health Psychology. 2010;29:374 – 383.

96. Neal DT, Wood W, Drolet A. How do people adhere to goals when willpower is low? The profits (and pitfalls) of strong habits. Journal of Personality and Social Psychology. 2013;104:959 – 975.

97. Lawson PJ, Flocke SA. Teachable moments for health behavior change: a concept analysis. Patient Educ Couns. 2009;76:25–30. doi: 10.1016/j.pec.2008.11.002

98. Rhodes R, de Bruijn GJ, Matheson DH. Habit in the physical activity domain: Integration with intention temporal stability and action control. Journal of Sport and Exercise Psychology. 2010;32:84 – 98.

99. Stroke Patient & Family Support Services. Stroke Warriors Group. NewYork-Presbyterian/Columbia University Irving Medical Center. https://www.neurology.columbia.edu/patient-care/specialties/stroke-and-cerebrovascular-disease/stroke-patient-family-support-services. Accessed 12/16/2024.

100. Lamont RA, Calitri R, Mounce LTA, Hollands L, Dean SG, Code C, Sanders A, Tarrant M. Shared social identity and perceived social support among stroke groups during the COVID-19 pandemic: Relationship with psychosocial health. Appl Psychol Health Well Being. 2023;15:172–192. doi: 10.1111/aphw.12348

